# Study Protocol: A Scoping Review on Costing Methodologies in Robotic Ventral Hernia Repair

**DOI:** 10.1101/2024.08.11.24311831

**Authors:** Kristian Als Nielsen, Karsten Kaiser, Per Helligsø, Michael Festersen Nielsen, Alexandros Valorenzos

## Abstract

**Background:** Healthcare expenditure is increasing at a rate that surpasses the growth of the gross domestic product in most Western nations, emphasizing the need for precise hospital accounting practices. In resource-constrained environments, accurately estimating the costs of hospital services, such as robotic ventral hernia repair, is essential for achieving efficiency and transparency. Despite the existence of several studies reporting on the cost of robotic ventral hernia repair, there is a lack of systematic mapping of current knowledge on the methodological designs used in studies reporting on the costs and resource use.

**Methods:** This study protocol outlines the methodology for a scoping review aimed at systematically mapping the existing literature on costing methodologies in robotic ventral hernia repair. The scoping review will follow the framework outlined by Arksey and O’Malley and adhere to the PRISMA-ScR guidelines. A systematic search will be conducted in Embase, Medline and Cochrane Library. Studies will be included if they involve patients undergoing robotic ventral hernia repair and report on cost/costing methodologies. The review will extract data on study characteristics, intervention specifics, and detailed costing methodologies. Two independent reviewers will conduct the data extraction, with discrepancies resolved through discussion or by a third reviewer.

**Discussion:** The review will identify significant variations in costing methodologies, including differences in perspectives (hospital vs. societal), time horizons, and cost components. It aims to highlight gaps and inconsistencies in the current literature, providing a foundation for future research to standardize costing methodologies and improve the accuracy of economic evaluations in robotic surgery. By systematically mapping the existing literature, this scoping review will provide valuable insights into the current state of costing methodologies in robotic ventral hernia repair. It will serve as a foundational reference for researchers, policymakers, and healthcare providers, offering recommendations to enhance the economic evaluation of robotic surgical technologies.

## Background

Healthcare expenditure is increasing at a rate that surpasses the growth of the gross domestic product in most Western European nations each year(1-3).This trend, coupled with global healthcare reforms, has intensified the dependence on precise hospital accounting practices. In environments with constrained resources, accurately estimating the costs of hospital services is essential for achieving efficiency and transparency. In many high-income countries, hospitals operate under Diagnosis Related Group (DRG)-based prospective payment systems, necessitating the identification and elimination of inefficiencies, particularly in services where production costs exceed pricing(4, 5). Consequently, hospitals require dependable patient-level cost estimates to accurately assess resource utilization. Detailed and pertinent cost information at the patient level is crucial for policymakers, payers, and healthcare providers.

However, accurately calculating costs within the hospital setting is challenging due to factors like case heterogeneity, labor intensity, and the complexity of production processes. Studies indicate significant cost variation for the same service, influenced by provider and patient characteristics, efficiency levels, clinical activities, and, critically, the costing method used(6). Previous systematic reviews comparing the cost-effectiveness of Robotic Minimally Invasive Surgery (RMIS) to other surgical methods have generally found RMIS to be more expensive(7). These conclusions, however, were drawn from limited evidence characterized by diverse study designs, methodologies, and follow-up durations.

Despite the existence of several articles that include an economic evaluation of robotic ventral hernia repair, there is a lack of systematic mapping of current knowledge on the methodological designs used in studies reporting on the costs and resource use of robotic surgery for ventral hernia repair. This scoping review aims to systematically describe the methodological designs employed in studies of resource use and costs related to robotic ventral hernia repair. The primary focus is not on comparing the cost of robotic surgery with other surgical methods, but on evaluating the various methodological choices that could impact the validity of cost analyses.

Conducting a scoping review of the literature on the costs and costing methodologies of robotic ventral hernia repair can enhance our understanding of the topic, including the different approaches used and the limitations and challenges associated with these methodologies. The objectives of this paper are to provide an overview of the existing literature on the costs of robotic ventral hernia repair, to examine the methodologies employed, to identify gaps in the research, and to offer recommendations for future research in this field.

## Methods and Design

The scoping review will be conducted following the methodology outlined by Arksey and O’Malley and adhering to the guidelines of the Preferred Reporting Items for Systematic Reviews and Meta-Analyses – Scoping Review extension (PRISMA-ScR). The study will be developed in the following five stages:

### Stage 1: Identification of the Research Question

This scoping review aims to answer the following questions:

- What is known from the existing literature about the cost of ventral hernia repair using a robotic platform?
- What are the methodological designs employed in studies reporting resource use and costs related to robotic ventral hernia repair, and how do these methodological choices impact the validity of cost analyses?

### Stage 2: Identification of Relevant Literature

A systematic search will be conducted in cooperation with a medical librarian from our institution using the following databases:

- Embase
- Medline
- Cochrane Library
- Scopus for citation tracking

An example of the search strategy for Embase (Ovid) is presented in Table 1. The search strategy will be translated for Medline and Cochrane Library.

**Table 1.** Embase (Ovid) search strategy.

|  |  |  |
| --- | --- | --- |
| Ventral hernia<br>block | 1 | abdominal wall hernia/ |
|  | 2 | umbilical hernia/ |
|  | 3 | incisional hernia/ |
|  | 4 | herni*.ti,ab,kf. |
| RMIS block | 5 | robot/ |
|  | 6 | robot*.ti,ab,kf. |
|  | 7 | robotics/ |
|  | 8 | robot assisted surgery/ |
|  | 9 | computer assisted surgery/ |
| Costing block | 10 | "cost"/ |
|  | 11 | cost*.ti,ab,kf. |
|  | 12 | economics/ |
|  | 13 | economics*.ti,ab,kf. |
|  | 14 | financial.ti,ab,kf. |
|  | 15 | pric*.ti,ab,kf. |
| Combination | 16 | 1 or 2 or 3 or 4 |
|  | 17 | 5 or 6 or 7 or 8 or 9 |
|  | 18 | 10 or 11 or 12 or 13 or 14 or 15 |
|  | 19 | 16 and 17 and 18 |

### Stage 3: Study Selection

Inclusion and exclusion criteria in accordance with the PCC principle are outlined below. Only peer-reviewed original research concerning costs in robotic ventral hernia repair will be included. Only English-language articles will be included. All results will be imported into Covidence for screening. All duplicates will be removed. Two reviewers will screen titles and abstracts using the inclusion and exclusion criteria outlined below. The same two reviewers will examine the remaining articles and select studies according to the inclusion criteria. Disagreements will be resolved through discussion and overseen by a third-party investigator. A PRISMA flowchart will be presented to summarize the search, screening, exclusion, and inclusion process for the study selection of relevant studies.

#### Inclusion Criteria

##### Population

- Studies involving patients undergoing robotic ventral hernia repair.

##### Concept

- Studies that report on cost and/or costing methodologies.

##### Context

- Studies conducted in hospitals or healthcare institutions where robotic ventral hernia repair is performed.
- Original research of all types, including randomized controlled trials, prospective and retrospective cohort studies, case-control studies, reviews, and economic evaluations.
- Studies published in any country and in English.

#### Exclusion Criteria

##### Population

- Studies not specifically addressing patients undergoing robotic ventral hernia repair.

##### Concept

- Studies that do not report details on costs and/or costing methodologies.

##### Context

- Non-English literature
- Grey literature

### Stage 4: Charting the Data

For this scoping review, a standardized data extraction form will be used to collect relevant details from each included study. This form will capture essential elements such as study characteristics (e.g., author, year, country, study design, and type of economic evaluation), intervention specifics (e.g., type of robotic ventral hernia repair and comparator techniques), and methodological aspects related to costing (e.g., perspective, time horizon, cost components, and calculation methods).

#### Specific Aspects to be Captured

1. Type of Economic Evaluation:
  - Cost minimization analysis
  - Cost-benefit framework
  - Activity-based costing
  - Cost studies
2. Costing Methodologies:
  - Microcosting
  - Gross costing
  - Hospital charges
3. Specific Costs of Relevance:
  - Purchasing Cost of the Robot:
    - Depreciation accounted for
    - Number of years the robot is expected to operate
    - Number of procedures per year the robot is used for
  - Maintenance of the Robot:
    - Service agreement costs
  - Preoperative Preparation of the Patients
  - Staff Salary:
    - Specific staff members (e.g., surgeons, ER nurses, anesthesiologists)
  - Surgical Equipment
  - Operating Room Costs:
    - Sterilization of equipment
    - Cleaning costs
  - Drug Costs:
    - Anesthesia
    - Pain medications
    - IV infusions
    - Antibiotics
  - Hospital Room Costs:
    - Level of detail (e.g., food for patients, cleaning, clothing)
    - Accounting for length of hospital stay
  - Post-Discharge Costs:
    - Readmission costs
    - Reoperation costs
    - Use of equipment (e.g., splints)
    - Pain medications
    - Outpatient control costs
4. Adjustments:
  - Whether the study accounts for the learning curve of using the robotic system
  - Whether the study accounts for the operation time
5. Perspective of Costing:
  - Hospital perspective
  - Societal perspective

Additionally, any reported challenges and limitations in the costing methodologies will be documented. The data charting process will be conducted by two independent reviewers, blinded to each other to ensure accuracy and comprehensiveness. Discrepancies will be resolved through discussion or by consulting a third reviewer.

This detailed approach to data charting will enable a thorough and nuanced understanding of the costing methodologies used in studies of robotic ventral hernia repair, and will help to identify gaps and inconsistencies in the literature.

### Stage 5: Collating, Summarizing, and Reporting of Results

The collected data will be systematically collated, summarized, and reported to provide a comprehensive overview of the findings. This process involves organizing the extracted data into meaningful categories that align with the research questions and objectives. Quantitative data will be presented in tabular and graphical formats to illustrate trends, frequencies, and distributions of costing methodologies used in robotic ventral hernia repair studies. This stage will also involve identifying patterns, inconsistencies, and gaps in the literature. The final report will present a detailed summary of the reviewed studies, highlighting key findings, methodological variations, and areas requiring further research. The results will be discussed in the context of existing knowledge, and recommendations will be made for future studies to enhance the understanding and accuracy of costing methodologies in robotic ventral hernia repair.

## Discussion

This scoping review aims to systematically map the existing literature on costs and costing methodologies in robotic ventral hernia repair. By including a wide range of studies, we intend to provide a comprehensive overview of the costs associated with the robotic platform in ventral hernia repair, how these costs are calculated, the methodological choices made, and the challenges encountered in this field. The review will highlight significant variations in costing methodologies, including differences in perspectives, time horizons, and cost components included in the analyses. Such variations can profoundly impact the reported cost-effectiveness of robotic surgery, potentially leading to inconsistent conclusions and impeding informed decision-making.

One of the anticipated findings is the lack of standardization in costing methodologies. The review is likely to reveal that many studies do not adhere to best practices in economic evaluations, such as including comprehensive cost components like the purchase and maintenance of robotic systems or adequately accounting for indirect costs. These gaps can undermine the validity and comparability of cost analyses, suggesting a need for more rigorous methodological standards.

Additionally, this review will identify key areas for future research, such as the need for longitudinal studies to capture long-term costs and benefits, and the importance of incorporating broader economic perspectives. By synthesizing the existing evidence, this review will provide valuable insights that can guide future research efforts and improve the transparency and reliability of costing studies in robotic ventral hernia repair.

Overall, this scoping review will serve as a foundational reference for researchers, policymakers, and healthcare providers, highlighting current knowledge, identifying gaps, and suggesting directions for future research to enhance the economic evaluation of robotic surgical technologies.

## Data Availability

No datasets were generated or analysed during the current study. All relevant data from this study will be made available upon study completion.

## Acknowledgements

We would like to express our sincere gratitude to Caroline Moos, Medical Librarian at the Department of Medical Research, University of Southern Denmark, for her invaluable guidance regarding systematic search strategies and methodology. Her expertise and support were instrumental in the development of this study protocol.

